# Effectiveness of mRNA COVID-19 vaccines and hybrid immunity in preventing SARS-CoV-2 infection and symptomatic COVID-19 among adults in the United States

**DOI:** 10.1101/2024.11.01.24316597

**Authors:** Leora R. Feldstein, Jasmine Ruffin, Ryan E. Wiegand, Jade James-Gist, Tara M. Babu, Craig B. Borkowf, Melissa Briggs-Hagen, James Chappell, Helen Y. Chu, Janet A. Englund, Jennifer L. Kuntz, Adam S. Lauring, Natalie Lo, Marco Carone, Christina Lockwood, Emily T. Martin, Claire M. Midgley, Arnold S. Monto, Allison L. Naleway, Tara Ogilvie, Sharon Saydah, Mark A. Schmidt, Jonathan E. Schmitz, Ning Smith, Ine Sohn, Lea Starita, H. Keipp Talbot, Ana A. Weil, Carlos G. Grijalva

## Abstract

**Background:** Understanding protection against SARS-CoV-2 infection by vaccine and hybrid immunity is important for informing public health strategies as new variants emerge.

**Methods:** We analyzed data from three cohort studies spanning September 1, 2022–July 31, 2023, to estimate COVID-19 vaccine effectiveness (VE) against SARS-CoV-2 infection and symptomatic COVID-19 among adults with and without prior infection in the United States. Participants collected weekly nasal swabs, irrespective of symptoms, annual blood draws, and completed periodic surveys, which included vaccination status and prior infection history. Swabs were tested molecularly for SARS-CoV-2. VE was estimated using Cox proportional hazards models for the hazard ratios of infections, adjusting for covariates. VE was calculated considering prior infection and recency of vaccination.

**Results:** Among 3,343 adults, adjusted VE of bivalent vaccine against infection was 37.2% (95% CI: 11.4-58.5%) within 7-60 days of vaccination and 17.0% (95% CI: -3.7-33.2%) within 60-179 days of vaccination compared to participants who were unvaccinated/received an original monovalent vaccine dose ≥180 days prior. Overall, adjusted VE of bivalent vaccine, in conjunction with prior infection, was 62.2% (95% CI: 44.2-74.6%) within 7-179 days of vaccination and 39.4% (95% CI: 11.7-61.3%) ≥180 days compared to naïve participants who were unvaccinated/received a monovalent vaccine dose ≥180 days prior.

**Conclusions:** Adults with both prior infection and recent vaccination had high protection against infection and symptomatic illness. Recent vaccination alone provided moderate protection.

## Introduction

Adults are most severely impacted by COVID-19 illness; more than 1.2 million COVID-19-related deaths among Americans aged ≥18 years have occurred as of October 28, 2024, accounting for 99.8% of all COVID-19-related deaths in the United States [1]. The original monovalent mRNA COVID-19 vaccines were highly effective at reducing risk of severe illness and death, but waned over time, especially for less severe outcomes, and effectiveness appeared lower against Omicron [2, 3]. To address the diminished protection from vaccination, the Food and Drug Administration (FDA) authorized use of the bivalent mRNA COVID-19 vaccine, composed of ancestral and Omicron BA.4/BA.5 spike proteins [4]. While previous studies have shown that bivalent mRNA COVID-19 vaccination among adults is effective at reducing COVID-19-related hospitalizations and death [5-7], fewer studies have assessed whether updated vaccines provide protection against infection and milder symptomatic illness [8-15], and examined the impact of prior infection in combination with receipt of the vaccine [8-10, 16, 17].

Understanding how well adults are protected against SARS-CoV-2 infection by vaccine alone and by hybrid immunity is important for informing public health strategies and policies, particularly as new variants continue to emerge. During a period of Omicron XBB variant predominance, this analysis used data from three prospective cohort studies to estimate effectiveness of authorized monovalent and bivalent COVID-19 vaccines (excluding the 2023-2024 monovalent vaccine) and history of prior infection against laboratory-confirmed SARS-CoV-2 virus infection and symptomatic COVID-19 among adults in the United States.

## Methods

### Study population

We conducted an analysis on data spanning September 1, 2022 – July 31, 2023 from four sites in the United States to estimate COVID-19 vaccine effectiveness (VE) among adults aged ≥18 years. Specifically, we combined data from three prospective cohort studies, CASCADIA, CoVE (Community Vaccine Effectiveness against Asymptomatic and Symptomatic SARS-CoV-2 Infection in Michigan), and VIEW (Viruses and Infections in Essential Workers) [18, 19]. CASCADIA enrolled Kaiser Permanente Northwest and University of Washington patients and community members in the Portland, Oregon and Seattle, Washington metropolitan areas (both children and adults aged 18-49). CoVE enrolled children and adults of all ages who live in Michigan and receive healthcare and VIEW enrolled adults who are essential (non-healthcare) workers in Tennessee.

For this study, adults living in Washington, Oregon, Michigan, and Tennessee, including individuals from the same household, were eligible for inclusion. Written informed consent was obtained from all participants. This study was reviewed by the US Centers for Disease Control and Prevention and approved by the institutional review boards at participating sites and was conducted consistent with applicable federal law and CDC policy [20].

### Data and specimen collection

At enrollment, participants completed an enrollment survey that included demographics, household characteristics, chronic medical conditions, COVID-19 vaccination history, and prior SARS-CoV-2 infection history; participants were resurveyed at regular intervals to capture up-to-date demographic information. Blood specimens were collected near to the time of enrollment from participants who consented to phlebotomy or by self-collection of blood specimens using Mitra or Tasso+ devices [21]. Weekly surveillance was conducted for COVID-like illness symptoms. Participants were asked to self-collect nasal swabs weekly, irrespective of symptoms. To optimally capture symptomatic COVID-19, participants were instructed to collect an additional respiratory specimen upon onset of symptoms if they occurred outside of the timing of their regular weekly swab cadence.

### Laboratory Testing

All respiratory specimens were tested for SARS-CoV-2 using real-time *reverse transcription-polymerase chain reaction* (RT-PCR) based assays, as detailed in the supplementary materials (eTable 1). Of note, while the component studies (CASCADIA, CoVE, VIEW) do not fall under the medicolegal auspices of *clinical* testing, all three utilized the same molecular assays employed for patient care. Less than 1% of specimens generated PCR results that would be consider either ‘inconclusive’ or ‘failed’ under clinically validated parameters; for the purposes of the present analyses, these specimens were considered negative. Whole genome sequencing was attempted on all SARS-CoV-2-positive specimens with an adequate viral load [22-25].

Available serum specimens were tested for the presence of anti-nucleocapsid (N) IgG using a quantitative MesoScale Discovery (MSD) VPLEX assays (eTable 1). For the SARS-CoV-2 MSD assay, titers against the N protein were interpolated from a standard calibration curve provided by the manufacturer. Specimens below the lower limit of quantitation per assay insert were set to a value of half the lower limit. Per the assay insert, specimens were determined to have detectable anti-N IgG if they had a titer equal to or greater than 5,000 assigned units per mL (AU/mL).

### Variables of interest

COVID-19 vaccination status was captured from enrollment and weekly/monthly surveys (self-report), vaccine cards provided by the participant, and/or from queries of the state immunization information systems and electronic medical records (EMR), when available. Vaccination data included vaccination dates, number of doses, and manufacturer. Information from the EMR and state immunization information systems was used preferentially over self-reported information in the event that a participant did not report a history of vaccination.

Symptomatic COVID-19 was defined as those with a positive RT-PCR test and at least two COVID-like illness symptoms reported within seven days before or after the specimen collection date. The list of COVID-like illness symptoms varied by the cohort study (eTable 2).

Prior infection was defined as laboratory-confirmation of infection by RT-PCR from a study-collected specimen prior to the analytic period, positive anti-N SARS-CoV-2 antibody at enrollment, or self-report of infection prior to enrollment or September 1, 2022 (whichever occurred later). Time since prior infection was categorized as no prior infection, <4 months, 4 – <6 months, 6 – <12 months, ≥12 months. Dates of prior infection were imputed for 198 (5.9%) participants who only had serologic results and therefore did not have dates associated with prior infection. Imputation was done using results from linear regression models, in which baseline nucleocapsid blood draw date and numeric nucleocapsid values served as the predictors for the date of prior infection (among study participants with known prior infection dates) (eMethods).

### Statistical analysis

Descriptive statistics, comparing participants who became infected during the study period to participants who remained uninfected, included frequencies (proportions) for categorical variables and medians (interquartile ranges [IQRs]) for continuous variables. P-values were calculated using chi-square tests for categorical variables and Wilcoxon rank sum tests for continuous variables. The Andersen-Gill extension of the Cox proportional hazards model with time-varying vaccination status was used to estimate hazard ratios (HR) of first SARS-CoV-2 infections, comparing participants with receipt of a bivalent dose to participants who were either unvaccinated or had received the original monovalent vaccine ≥180 days prior [26]. Separate VE estimates were produced for SARS-CoV-2 infection (inclusive of asymptomatic and symptomatic infections), symptomatic COVID-19, vaccine valency (original monovalent vs. bivalent), and timing of vaccine receipt. VE estimates were stratified by prior infection status and variant period among those with recent bivalent vaccination (within 7-179 days). Additionally, VE estimates were produced for those with both vaccination and prior infection, with naïve (no evidence of prior infection) participants who were either unvaccinated or had received the original monovalent vaccine ≥180 days prior as the reference group.

Multivariable models adjusted for age, sex, race/ethnicity, presence of at least one underlying health conditions, time since prior infection, geographic site, household size, and 7-day average of COVID-19 cases per 100 000 by site (local incidence: modeled as a continuous linear variable). COVID-19 vaccine effectiveness (VE) was calculated as VE = (1 - HR) × 100. Confidence intervals were calculated using the standard estimation methods for the Cox proportional hazards model, because the cluster size of participants by household was small [27]. In particular, 35.4% of households had two or more adults included in the analysis (note: 89 participants had a missing household ID and were assumed to be the only study participant in the household).

Person-time was calculated as the total number of days under surveillance for a given vaccination status during the analytic period. The surveillance period started on September 1, 2022 and ended on the date of a participant’s first positive RT-PCR test, the participant’s study withdrawal date, or end of the analytic period (July 31, 2023), whichever came first. Individuals enrolled after September 1, 2022 began time at risk at the time of surveillance start or six weeks after prior infection, if recently infected prior to enrollment. The surveillance weeks for which there was no specimen result (e.g., participant skipped a weekly swab) for four or more consecutive weeks were censored. The two weeks following an original monovalent primary vaccine dose and the week following bivalent or original monovalent booster vaccine doses were also excluded from person-time. A sensitivity analysis was conducted by restricting the analysis period to November 27, 2022 – July 31, 2023 in order to account for the difference in enrollment start dates of the studies (e.g. CASCADIA and CoVE began enrollment in July and August of 2022, whereas VIEW started in November 2022) and for the differences in the percent of participants who received a bivalent vaccine by site (CASCADIA=68.2%, CoVE=59.9%, and VIEW=25.4%).

All analyses were conducted using SAS software (version 9.4; SAS Institute, Cary, NC) or R Studio software (version 4.1.0; R Foundation).

## Results

### Study population

Between September 1, 2022 and July 31, 2023, 3,343 participants contributed to person-time in the analysis, 50.0% from CASCADIA, 39.4% from VIEW, and 10.6% from CoVE (Table 1). Overall, 67.2% were female, median age was 41 years (interquartile range [IQR]: 36-46 years), and the majority were White, non-Hispanic (69.2%). Almost half of participants lived in a household with 4 or more individuals (48.1%), and 60.2% of participants reported having at least one chronic health condition. Age, gender, race/ethnicity, prevalence of chronic conditions, and household size varied by site (Table e3). Of the 1462 prior infections reported, 13.3% were from self-report only. During the study period, 21.7% (n=727) of participants had a laboratory-confirmed SARS-CoV-2 infection. A higher proportion of participants living in a household with more than one other person had SARS-CoV-2 infections than those who lived alone (20.6% versus 14.2%). A higher proportion of those with no documented prior infection had SARS-CoV-2 infection during the study period than those with a prior infection (26.2% versus 16.1%). Among participants with SARS-CoV-2 infections during the study period, 62.3% reported symptomatic COVID-19. Of the 727 SARS-CoV-2 infections, 400 (55%) had genetic sequencing results; the most prevalent lineages were XBB (60.3%), BQ.1.1 (14.8%), and BA.4/BA.5 (14.0%).

**Table 1.** Characteristics of participants by laboratory-confirmed SARS-CoV-2 status, September 1, 2022 – July 31, 2023.

|  | Overall<br>No. (Col %) |  | SARS-CoV-2<br>positive during the<br>study period<br>No. (Row %) |  | SARS-CoV-2<br>negative<br>No. (Row %) |  | P-value<sup>a</sup> |
| --- | --- | --- | --- | --- | --- | --- | --- |
| **Total** | 3343 |  | 727 | 21.7% | 2616 | 78.3% |  |
| **Site** |  |  |  |  |  |  | 0.258 |
| CASCADIA: Kaiser Permanente Northwest | 854 | 25.5% | 186 | 21.8% | 668 | 78.2% |  |
| CASCADIA: University of Washington | 819 | 24.5% | 197 | 24.1% | 622 | 75.9% |  |
| CoVE: Michigan | 354 | 10.6% | 70 | 19.8% | 284 | 80.2% |  |
| VIEW: Tennessee | 1316 | 39.4% | 274 | 20.8% | 1042 | 79.2% |  |
| **Sex** |  |  |  |  |  |  | 0.985 |
| Female | 2247 | 67.2% | 489 | 21.8% | 1758 | 78.2% |  |
| Male | 1086 | 32.5% | 237 | 21.8% | 849 | 78.2% |  |
| Non-female or male | 5 | 0.1% | 1 | 20.0% | 4 | 80.0% |  |
| **Age, median (IQR) (years)** | 41 | 36-46 | 41 | 37-46 | 41 | 36-47 | 0.645 |
| **Age group (years)** |  |  |  |  |  |  | 0.189 |
| 18-49 | 2880 | 86.2% | 641 | 22.3% | 2239 | 77.7% |  |
| 50-64 | 386 | 11.5% | 73 | 18.9% | 313 | 81.1% |  |
| 65+ | 77 | 2.3% | 13 | 16.9% | 64 | 83.1% |  |
| **Race/Ethnicity** |  |  |  |  |  |  | 0.030 |
| White, non-Hispanic | 2312 | 69.2% | 531 | 23.0% | 1781 | 77.0% |  |
| Hispanic or Latino | 252 | 7.5% | 59 | 23.4% | 193 | 76.6% |  |
| Multiple races, non-Hispanic | 127 | 3.8% | 23 | 18.1% | 104 | 81.9% |  |
| Black, non-Hispanic | 317 | 9.5% | 54 | 17.0% | 263 | 83.0% |  |
| Other, non-Hispanic<sup>b</sup> | 251 | 7.5% | 43 | 17.1% | 208 | 82.9% |  |
| **Chronic Conditions<sup>c</sup>** |  |  |  |  |  |  | 0.078 |
| None | 1331 | 39.8% | 310 | 23.3% | 1021 | 76.7% |  |
| 1 or more | 2012 | 60.2% | 417 | 20.7% | 1595 | 79.3% |  |
| **Individuals living in household** |  |  |  |  |  |  | 0.011 |
| 1 | 275 | 8.2% | 39 | 14.2% | 236 | 85.8% |  |
| 2 | 653 | 19.5% | 137 | 21.0% | 516 | 79.0% |  |
| 3 | 755 | 22.6% | 173 | 22.9% | 582 | 77.1% |  |
| ≥4 | 1642 | 49.1% | 375 | 22.8% | 1267 | 77.2% |  |
| **Weekly swab adherence (%), median (IQR)** | 85 | 74-93 | 86 | 76-93 | 85 | 73-93 | 0.051 |
| **Swab adherence** |  |  |  |  |  |  | 0.007 |
| <80% | 1248 | 37.3% | 240 | 19.2% | 1008 | 80.8% |  |
| ≥80% | 2095 | 62.7% | 487 | 23.2% | 1608 | 76.8% |  |
| **Prior infection<sup>d</sup>** |  |  |  |  |  |  | <0.001 |
| None | 1881 | 56.3% | 492 | 26.2% | 1389 | 73.8% |  |
| 1 or more | 1462 | 43.7% | 235 | 16.1% | 1227 | 83.9% |  |
| **Time since prior infection<sup>d,e</sup>** |  |  |  |  |  |  | <0.001 |
| No prior infection | 1881 | 56.3% | 492 | 26.2% | 1389 | 73.8% |  |
| <4 months | 485 | 14.5% | 72 | 14.8% | 417 | 86.0% |  |
| 4-<6 months | 299 | 8.9% | 40 | 13.4% | 259 | 86.6% |  |
| 6-<12 months | 375 | 11.2% | 62 | 16.5% | 313 | 83.5% |  |
| ≥12 months | 298 | 8.9% | 61 | 20.5% | 237 | 79.5% |  |
| **Symptomatic COVID-19<sup>f</sup>** |  |  |  |  |  |  |  |
| No | - | - | 274 | 37.7% | - | - |  |
| Yes | - | - | 453 | 62.3% | - | - |  |
| **Predominant variant period of infection<sup>g</sup>** |  |  |  |  |  |  |  |
| BA.4/BA.5<sup>h</sup> | - | - | 242 | 33.3% | - | - |  |
| XBB<sup>i</sup> | - | - | 485 | 66.7% | - | - |  |

Abbreviations: Col = column; IQR = interquartile range.
<sup>a</sup> Fisher's exact tests, Kruskal-Wallis rank sum test, and Pearson's Chi-squared tests were used to calculate p-values.
<sup>b</sup> The 'Other, non-Hispanic' category includes participations who identified as American Indian non-Hispanic, Alaska Native non-Hispanic, Asian non-Hispanic, Black and African American non-Hispanic, and Native Hawaiian/Pacific Islander non-Hispanic.
<sup>c</sup> Chronic conditions for CASCADIA and CoVE included: asthma, heart disease, sleep apnea, down syndrome, diabetes, cancer, autoimmune disease, liver disease, kidney disease, hematological disease, neurologic or neuromuscular disease, stroke, deep vein thrombosis or pulmonary embolism, anxiety, depression, immunosuppression, hypertension and thyroid disease. For VIEW: asthma, chronic pulmonary disease, obesity, heart disease, diabetes, liver disease, kidney disease, cancer, arthritis, hematological disease, neurologic or neuromuscular disease, stroke, deep vein thrombosis or pulmonary embolism, anxiety, depression, immunosuppression, hypertension and thyroid disease.
<sup>d</sup> Prior infection was defined as laboratory-confirmation of infection by RT-PCR from a study-collected specimen, positive anti-N SARS-CoV-2 antibody, or self-report of infection prior to enrollment or September 1, 2022.
<sup>e</sup> Time since prior infection was calculated as the date of the prior infection to the first week each participant was included in the analysis.
<sup>f</sup> Symptomatic COVID-19 was defined as those with a positive RT-PCR test and at least two COVID-like illness symptoms reported within seven days of the specimen collection date.
<sup>g</sup> Time period in which the positive SARS-CoV-2 infection occurred.
<sup>h</sup> BA.4/BA.5 predominant period was defined as September 1, 2022 – January 27, 2023.
<sup>i</sup> XBB predominant period was defined as January 28, 2023 – July 31, 2023.

### Vaccine uptake

Half of participants received at least one bivalent COVID-19 vaccine dose (49.7%) (Table 2). Participants enrolled from the Kaiser Permanent Northwest health plan (Oregon and Washington) had the highest uptake of bivalent vaccine doses (77.0%), whereas those in Tennessee (VIEW) had the lowest (25.4%). Black, non-Hispanic participants had the lowest reported proportion of receiving bivalent vaccine (27.8%), followed by Hispanic participants (34.5%), compared to White, non-Hispanic participants (54.1%). Those without report of a prior infection had higher uptake of bivalent vaccine (53.4%) compared to those with report of a prior infection (44.9%).

**Table 2.** Characteristics of participants by COVID-19 vaccination status, September 1, 2022 – July 31, 2023.

|  | Overall<br>No. (Col %) |  | Unvaccinated or<br>monovalent only<br>No. (Row %) |  | Bivalent dose<br>No. (Row %) |  | P-value<sup>a</sup> |
| --- | --- | --- | --- | --- | --- | --- | --- |
| **Total** | 3343 |  | 1682 | 50.3% | 1661 | 49.7% | <0.001 |
| **Site** |  |  |  |  |  |  |  |
| CASCADIA: Kaiser Permanente Northwest | 854 | 25.5% | 196 | 23.0% | 658 | 77.0% |  |
| CASCADIA: University of Washington | 819 | 24.5% | 362 | 44.2% | 457 | 55.8% |  |
| CoVE: Michigan | 354 | 10.6% | 142 | 40.1% | 212 | 59.9% |  |
| VIEW: Tennessee | 1316 | 39.4% | 982 | 74.6% | 334 | 25.4% |  |
| **Sex** |  |  |  |  |  |  | 0.297 |
| Female | 2247 | 67.2% | 1141 | 50.8% | 1106 | 49.2% |  |
| Male | 1086 | 32.5% | 536 | 49.4% | 550 | 50.6% |  |
| Other | 5 | 0.1% | 1 | 20.0% | 4 | 80.0% |  |
| **Age, median (IQR) (years)** | 41 | 36-46 | 40 | 34-46 | 42 | 38-47 | <0.001 |
| **Age group (years)** |  |  |  |  |  |  | <0.001 |
| 18-49 | 2880 | 86.2% | 1414 | 49.1% | 1466 | 50.9% |  |
| 50-64 | 386 | 11.5% | 234 | 60.6% | 152 | 39.4% |  |
| 65+ | 77 | 2.3% | 34 | 44.2% | 43 | 55.8% |  |
| **Race/Ethnicity** |  |  |  |  |  |  | <0.001 |
| White, non-Hispanic | 2312 | 69.2% | 1062 | 45.9% | 1250 | 54.1% |  |
| Hispanic or Latino | 252 | 7.5% | 165 | 65.5% | 87 | 34.5% |  |
| Multiple races, non-Hispanic | 127 | 3.8% | 56 | 44.1% | 71 | 55.9% |  |
| Black, non-Hispanic | 317 | 9.5% | 229 | 72.2% | 88 | 27.8% |  |
| Other, non-Hispanic<sup>b</sup> | 251 | 7.5% | 105 | 41.8% | 146 | 58.2% |  |
| **Chronic Conditions<sup>c</sup>** |  |  |  |  |  |  | 0.070 |
| None | 1331 | 39.8% | 644 | 48.4% | 687 | 51.6% |  |
| 1 or more | 2012 | 60.2% | 1038 | 51.6% | 974 | 48.4% |  |
| **Individuals living in household** |  |  |  |  |  |  | <.0001 |
| 1 | 275 | 8.2% | 158 | 57.5% | 117 | 42.5% |  |
| 2 | 653 | 19.5% | 402 | 61.6% | 251 | 38.4% |  |
| 3 | 755 | 22.6% | 349 | 46.2% | 406 | 53.8% |  |
| ≥4 | 1642 | 49.1% | 757 | 46.1% | 885 | 53.9% |  |
| **Weekly swab adherence (%), median (IQR)** | 85 | 74-93 | 81 | 70-92 | 88 | 78-95 | <0.001 |
| **Swab adherence** |  |  |  |  |  |  | <0.001 |
| <80% | 1248 | 37.3% | 778 | 62.3% | 470 | 37.7% |  |
| ≥80% | 2095 | 62.7% | 904 | 43.2% | 1191 | 56.8% |  |
| **Prior infection<sup>d</sup>** |  |  |  |  |  |  | <0.001 |
| None | 1881 | 56.3% | 877 | 46.6% | 1004 | 53.4% |  |
| 1 or more | 1462 | 43.7% | 805 | 55.1% | 657 | 44.9% |  |
| **Time since prior infection<sup>d,e</sup>** |  |  |  |  |  |  | <0.001 |
| No prior infection | 1881 | 56.3% | 877 | 46.6% | 1004 | 53.3% |  |
| <4 months | 485 | 14.5% | 223 | 46.0% | 262 | 54.0% |  |
| 4-<6 months | 299 | 8.9% | 166 | 55.5% | 133 | 44.5% |  |
| 6-<12 months | 375 | 11.2% | 198 | 52.8% | 177 | 47.2% |  |
| ≥12 months | 298 | 8.9% | 217 | 72.8% | 81 | 27.2% |  |
| **Symptomatic COVID-19<sup>f</sup>** |  |  |  |  |  |  | **<0.001** |
| No |  |  | 159 | 65.2% | 85 | 34.8% |  |
| Yes |  |  | 247 | 51.1% | 236 | 48.9% |  |

Abbreviations: Col = column; IQR = interquartile range.
<sup>a</sup>Fisher's exact tests, Kruskal-Wallis rank sum test, and Pearson's Chi-squared tests were used to calculate p-values.
<sup>b</sup>The 'Other, non-Hispanic' category includes participations who identified as American Indian non-Hispanic, Alaska Native non-Hispanic, Asian non-Hispanic, Black and African American non-Hispanic, and Native Hawaiian/Pacific Islander non-Hispanic.
<sup>c</sup>Chronic conditions for CASCADIA and CoVE included: asthma, heart disease, sleep apnea, down syndrome, diabetes, cancer, autoimmune disease, liver disease, kidney disease, hematological disease, neurologic or neuromuscular disease, stroke, deep vein thrombosis or pulmonary embolism, anxiety, depression, immunosuppression, hypertension and thyroid disease. For VIEW: asthma, chronic pulmonary disease, obesity, heart disease, diabetes, liver disease, kidney disease, cancer, arthritis, hematological disease, neurologic or neuromuscular disease, stroke, deep vein thrombosis or pulmonary embolism, anxiety, depression, immunosuppression, hypertension and thyroid disease.
<sup>d</sup>Prior infection was defined as laboratory-confirmation of infection by RT-PCR from a study-collected specimen, positive anti-N SARS-CoV-2 antibody, or self-report of infection prior to enrollment or September 1, 2022.
<sup>e</sup>Time since prior infection was calculated as the date of the prior infection to the first week each participant was included in the analysis.
<sup>f</sup>Symptomatic COVID-19 was defined as those with a positive RT-PCR test and at least two COVID-like illness symptoms reported within seven days of the specimen collection date.

### Vaccine effectiveness against infection

Of the 727 SARS-CoV-2 infections, 406 (55.8%) were among participants who were either unvaccinated/received a monovalent vaccine dose ≥180 days prior (1.74 infections per 1,000 person-days, [95% CI: (1.56–1.93]), and 321 (44.2%) were among those who received a bivalent dose (1.20 infections per 1,000 person-days, [95% CI: (1.08–1.34]) (Table 2 and 3). Adjusted VE of a bivalent dose received within 7-60 days against laboratory-confirmed SARS-CoV-2 infection, compared to the reference of being unvaccinated/receiving an original monovalent vaccine dose ≥180 days prior was 37.2% (95% CI: 11.4–58.5%) (Table 3). Compared to the same reference, adjusted VE of a bivalent dose received within 60-179 days was 17.0% (95% CI: -3.7-33.2%), and adjusted VE of a bivalent dose received ≥180 days prior was 8.7% (95% CI: -16.3-30.6%). When stratified by prior infection status, adjusted VE of a bivalent dose within 7-179 days against infection was 26.5% (95% CI: -1.1-40.8) among those who were naïve and 37.7% (95% CI: 6.8-58.2%) among those with prior infection. Adjusted VE of the original monovalent vaccine within 180 days against laboratory-confirmed SARS-CoV-2 infection, compared to the same reference group, was 26.7% (95% CI: -0.7-44.0%).

**Table 3.** COVID-19 vaccine effectiveness against laboratory-confirmed SARS-CoV-2 infection among adults by vaccine type, interval since receipt of dose, prior infection status, and variant period.

|  | Contributing participants<sup>a</sup> | Median observation day from vaccination<sup>b</sup> (IQR) | SARS-CoV-2 infections | Crude incidence rate of SARS-CoV-2 infections per 1000 PD | Unadjusted VE (95% CI) | Adjusted VE against infection (95% CI) |
| --- | --- | --- | --- | --- | --- | --- |
| **Interval since receipt of dose** |  |  |  |  |  |  |
| Unvaccinated or monovalent vaccine ≥180 days | 2013 | 446 (311, 569) | 352 | 1.74 (1.56, 1.93) | Ref | Ref |
| Monovalent vaccine, <180 days ago | 435 | 98 (55, 140) | 68 | 1.42 (1.11, 1.80) | 37.9 (17.3, 53.4) | 26.7 (-0.7, 44.0) |
| Bivalent vaccine, 7-60 days<sup>c</sup> | 859 | 35 (22, 47.8) | 40 | 1.08 (0.77, 1.47) | 48.1 (26.5, 63.3) | 37.2 (11.4, 58.5) |
| Bivalent vaccine, 60-179 days<sup>c</sup> | 1380 | 103.5 (61, 143) | 206 | 1.38 (1.20, 1.59) | 37.3 (21.0, 50.2) | 17.0 (-3.7, 33.2) |
| Bivalent vaccine, ≥180 days<sup>c</sup> | 1356 | 236 (208, 267) | 119 | 0.99 (0.82, 1.19) | 22.5 (1.5, 39.1) | 8.7 (-16.3, 30.6) |
| Bivalent vaccine, overall | 1695 | 165 (95, 228) | 327 | 1.20 (1.08, 1.34) | 34.7 (22.7, 44.9) | 18.5 (1.7, 32.2) |
| **Prior infection status** |  |  |  |  |  |  |
| Naïve, unvaccinated or monovalent vaccine ≥180 days<sup>d</sup> | 1040 | 431 (294, 563) | 199 | 2.02 (1.75, 2.32) | Ref | Ref |
| Naïve, bivalent vaccine within 7-179 days<sup>d</sup> | 835 | 104 (60, 143) | 163 | 1.90 (1.62, 2.22) | 30.9 (11.8, 45.8) | 26.5 (-1.1, 40.8) |
| Prior infection, unvaccinated or monovalent vaccine ≥180 days<sup>e</sup> | 973 | 458 (334, 575) | 153 | 1.48 (1.25, 1.73) | Ref | Ref |
| Prior infection, bivalent vaccine within 7-179 days<sup>e</sup> | 566 | 103 (61, 143) | 45 | 0.68 (0.50, 0.91) | 65.9 (51.1, 76.3) | 37.7 (6.8, 58.2) |

Abbreviations: IQR = interquartile range; PD = person-days; Ref = referent group; VE = vaccine effectiveness.
<sup>a</sup>Contributing participants in vaccination categories do not equal the number of participants in the study because participants could contribute to more than one vaccination category since vaccination status is time-varying.
<sup>b</sup>Adjusted estimates control for coefficient estimates of age, sex, race/ethnicity, underlying health conditions, time since prior infection, geographic site, household size, and 7-day average of COVID-19 cases per 100 000 by site.
<sup>c</sup>Data limited to October 30, 2022, to January 31, 2023, to allow both categories to possess the same amount of calendar time at risk.
<sup>d</sup>Naïve is defined as no evidence of prior infection before September 1, 2022. Prior infection was defined as laboratory-confirmation of infection by RT-PCR from a study-collected specimen, positive anti-N SARS-CoV-2 antibody, or self-report of infection prior to enrollment or September 1, 2022 (whichever occurred later).
<sup>e</sup>Prior infection was defined as laboratory-confirmation of infection by RT-PCR from a study-collected specimen, positive anti-N SARS-CoV-2 antibody, or self-report of infection prior to enrollment or September 1, 2022 (whichever occurred later).

### Hybrid immunity against infection and symptomatic illness

The combined protection from bivalent vaccination and prior infection compared to naïve participants who were unvaccinated/received a monovalent vaccine dose ≥180 days prior was 62.2 (95% CI: 44.2-74.6%) when vaccination was received within 7-179 days and 39.4% (95% CI: 11.7-61.3%) when received ≥180 days prior (Table 4). For symptomatic COVID-19 illness, combined protection was 73.0% (95% CI: 57.5-83.9%) when bivalent vaccination was received within 7-179 days and 56.7% (95% CI: 29.7-77.3%) when received ≥180 days prior.

**Table 4.** Hybrid protection against laboratory-confirmed SARS-CoV-2 infection among adults by vaccine type and interval since receipt of dose.

| Vaccine type, interval since receipt of vaccine, and prior infection status | Contributing participants<sup>a</sup> | Median observation day from vaccination<sup>b</sup> (IQR) | SARS-CoV-2 infections | Crude incidence rate of SARS-CoV-2 infections per 1000 PD | Unadjusted VE (95% CI) | Adjusted VE against infection<sup>c</sup> (95% CI) | Symptomatic SARS-CoV-2 infections<sup>d</sup> | Crude incidence rate of symptomatic COVID-19 per 1000 PD | Unadjusted VE against symptomatic COVID-19 (95% CI) | Adjusted VE against symptomatic COVID-19 (95% CI) |
| --- | --- | --- | --- | --- | --- | --- | --- | --- | --- | --- |
| Naive<sup>c</sup> & unvaccinated or monovalent vaccine ≥180 days | 1040 | 431 (294, 563) | 199 | 2.02 (1.75, 2.32) | Ref | Ref | 117 | 1.19 (0.98, 1.42) | Ref | Ref |
| Prior infection<sup>d</sup> & monovalent vaccine ≥180 days | 870 | 458 (334, 575) | 133 | 1.44 (1.21, 1.71) | 29.2 (11.1, 43.6) | 28.7 (10.3, 43.1) | 48 | 0.52 (0.38, 0.69) | 56.2 (37.6, 69.3) | 56.4 (40.3, 70.7) |
| Prior infection<sup>d</sup> & bivalent vaccine ≥180 days | 571 | 233 (206, 264) | 37 | 0.75 (0.53, 1.04) | 50.7 (27.8, 66.3) | 39.4 (11.7, 61.3) | 21 | 0.43 (0.27, 0.65) | 52.9 (20.6, 72.1) | 56.7 (29.7, 77.3) |
| Prior infection<sup>d</sup> & monovalent vaccine <180 days | 150 | 101 (57, 141) | 15 | 0.87 (0.49, 1.44) | 66.5 (41.7, 80.8) | 59.5 (31.5, 80.4) | 6 | 0.35 (0.13, 0.76) | 76.0 (44.7, 89.6) | 80.6 (59.2, 94.1) |
| Prior infection<sup>d</sup> & bivalent vaccine <180 days | 566 | 103 (61, 143) | 45 | 0.68 (0.50, 0.91) | 74.2 (63.6, 81.7) | 62.2 (44.2, 74.6) | 25 | 0.38 (0.25, 0.56) | 72.6 (56.6, 82.7) | 73.0 (57.5, 83.9) |

Abbreviations: IQR = interquartile range; PD = person-days; Ref = referent group; VE = vaccine effectiveness.
<sup>a</sup>Contributing participants in vaccination categories do not equal the number of participants in the study because participants could contribute to more than one vaccination category since vaccination status is time-varying.
<sup>b</sup>Adjusted estimates control for coefficient estimates of age, sex, race/ethnicity, underlying health conditions, prior infection status, geographic site, household size, and 7-day average of COVID-19 cases per 100 000 by site.
<sup>c</sup>Naive is defined as no evidence of prior infection before September 1, 2022.
<sup>d</sup>Prior infection was defined as laboratory-confirmation of infection by RT-PCR from a study-collected specimen, positive anti-N SARS-CoV-2 antibody, or self-report of infection prior to enrollment or September 1, 2022 (whichever occurred later).

The combined protection from monovalent vaccination within 7-179 days and prior infection compared to the same referenced group was 59.5% (95% CI: 31.5-80.4%) against laboratory-confirmed SARS-CoV-2 infection and 80.6% (95% CI: 59.2-94.1%) against symptomatic COVID-19 illness.

### Sensitivity analysis

In a sensitivity analysis, limiting the analysis timeframe to November 27, 2022 – July 31, 2023, adjusted VE of bivalent dose received within 7-60 days against infection was 46.2% (95% CI: 17.5–66.3%), received within 7-180 days was 26.9% (95% CI: 9.6–40.6%), and received ≥180 days was 8.5% (95% CI: - 19.0–31.8%) (Table e4). Overall, adjusted VE of a bivalent dose, regardless of timing of vaccine receipt, was 19.9% (95% CI: 3.2–33.2%). When examining protection from both vaccination and prior infection, protection against infection and symptomatic COVID-19 was similar to the main analysis results (Table e5).

## Discussion

COVID-19 vaccines have been shown to reduce the risk of severe illness and healthcare utilization following SARS-CoV-2 infection, but there are less data available on the impact of vaccination on the overall risk of infection [5-7]. In this multistate prospective community cohort study, adults who were vaccinated with a bivalent mRNA COVID-19 vaccine within the past 60 days were less likely to be infected with SARS-CoV-2 infection than those who were unvaccinated or received a monovalent vaccine dose ≥180 days prior regardless of prior infection history; overall adjusted VE of a bivalent dose was 37% within 7-60 days of receipt. Adults vaccinated >60 days prior had no measurable protection against infection. When hybrid immunity was evaluated, we found that adults with evidence of a prior infection and receipt of a COVID-19 vaccine within 7-179 days, regardless of valency, were less likely to be infected with SARS-CoV-2 infection and experience symptomatic COVID-19 illness than naïve individuals who were either unvaccinated or had received a monovalent vaccine ≥180 days prior; overall protection was estimated to be 60%-62% against infection and 73%-81% against symptomatic illness. In contrast, among adults with no evidence of prior infection, VE was lower (27% against infection when vaccination was received within 7-180 days), albeit this estimate was less precise due to limited power.

These findings suggest that hybrid immunity provided the strongest protection against SARS-CoV-2 infection, with the bivalent vaccine alone providing some protection against SARS-CoV-2 infection; however, vaccine effectiveness waned more rapidly than hybrid immunity and after 6 months there was no measurable protection. Our VE results are consistent with previous bivalent vaccine VE estimates against infection reported from other settings and some of these studies also found evidence of waning vaccine effectiveness against infection [8-10, 13, 15, 28]. Decreased protection over time may reflect either waning immunity from the vaccine and/or lower effectiveness of the vaccine against newly circulating variants or subvariants, such as XBB, which constituted the majority of infections in this analysis [3, 29]. Adults with documented prior infection had greater and more durable protection from both the original monovalent and bivalent vaccines within 179 days of receipt, which suggests that hybrid immunity against SARS-CoV-2 infection provides better protection than vaccination alone. These findings are also consistent with other studies that showed enhanced and longer-lasting protection against infection and symptomatic illness among individuals with a history both prior infection and vaccination against infection and symptomatic illness, but with diminishing protection over time [10, 16, 17].

### Limitations

There are several important limitations of this study. First, RT-PCR testing methods and COVID-like illness definitions varied by cohort site; therefore, some differences in definition of infection or symptomatic COVID-19 may be present. Second, weekly or symptomatic RT-PCR testing prior to the analytic study start date for estimation of prior infection history was only available among a subset of participants. To address this concern, we incorporated serologic data in order to identify additional prior SARS-CoV-2 infections but due to anti-N SARS-CoV-2 antibody waning, some prior infections may have been undetected. Third, social desirability or recall bias may have affected self-report of prior infection and vaccination status when RT-PCR and serologic test results and data from the state immunization information systems and EMR were unavailable. Fourth, vaccination may be associated with other protective factors that may be difficult to ascertain and account for fully. Fifth, limited sample sizes resulted in imprecise VE estimates and should be interpreted with caution, as the imprecision may indicate that the actual VE could be substantially different from the point estimates shown. Last, these observations derived from three large prospective cohort studies, while internally valid, may not directly generalize to other settings.

This study also has many strengths, including more than 3,300 participants enrolled from four distinct sites in the U.S. Participants swabbed weekly, regardless of symptoms, which greatly reducing the risk of missing an asymptomatic SARS-CoV-2 infection. Furthermore, adherence to weekly swabbing was high (median: 85%). Weekly and quarterly surveys, as well as data from the state immunization information systems and EMR, ensured detailed and complete information on potential confounding variables and vaccination status.

## Conclusion

Findings from this study demonstrate that during an Omicron predominant period, hybrid immunity provided the strongest protection against SARS-CoV-2 infection and symptomatic COVID-19. The bivalent COVID-19 vaccine also provided some protection. Protection from both were substantially lower 180 days or more following vaccination. Remaining up to date with recommended COVID-19 vaccinations and timing the receipt of vaccination shortly before peak respiratory virus season (presuming SARS-CoV-2 circulations adopts this typical pattern) may reduce SARS-CoV-2 infections.

## Data Availability

All data produced in the present analysis are restricted until the completion of the study.

## Funding

This study was supported by the National Center for Immunization and Respiratory Diseases, Centers for Disease Control and Prevention under contract numbers: 75D30121C12297 to Kaiser Foundation Hospitals, 75D30122C13149 to the University of Michigan, and 75D30122C13379 to Vanderbilt University. This project has also been funded in part with Federal funds from the National Institute of Allergy and Infectious Diseases, National Institutes of Health, Department of Health, and Human Services, under Contract No. 75N93021C00015. The Vanderbilt Institute for Clinical and Translational Research (VICTR) is funded by the National Center for Advancing Translational Sciences (NCATS) Clinical Translational Science Award (CTSA) Program, Award Number 5UL1TR002243-03. The content is solely the responsibility of the authors and does not necessarily represent the official views of the NIH.

### Disclaimer

*The findings and conclusions in this manuscript are those of the authors and do not necessarily represent the official position of the Centers for Disease Control and Prevention*.

## Acknowledgements

**Kaiser Permanente Center for Health Research:** Michael Allison, Deralyn Almaguer, David Amy, Britt Ash, Kristi Bays, Tara Beatty, Kristin Bialobok, Allison Bianchi, Cassandra Boisvert, Cathleen Bourdoin, Delanie Brown, Stacy Bunnell, Joseph Cerizo, Evelin Coto, Phil Crawford, Robin Daily, Lantoria Davis, Kristin Delaney, Stephen Fortmann, Lisa Fox, Kendall Frimodig, Kenni Graham, Holly Groom, Tarika Holness, Matt Hornbrook, Serah Kimachia, Emily Jubitz, Terry Kimes, Keelee Kloer, Dorothy Kurdyla, Isaiah Lankham, Teri Lawer, Caroline Lee, Max Lin, Richard Martin, Bryony Melcher, Richard Mularski, John Ogden, Chester Pabiniak, Aaron Piepert, Joanne Price, Sacha Reich, Angela Reyes-Ochoa, Jennifer Rivelli, Sperry Robinson, Katrina Schell, Emily Schield, Meagan Shaw, Anna Shivinsky, Nina Shockman, Ellen Sullivan, Martin Simer, Valencia Smith, Senait Tadesse, Alexandra Varga, Meredith Vandermeer, Brooke Wainwright, Mica Werner, Danika Whitcomb, Neil Yetz, Rebecca Ziebell; **University of Michigan:** Joshua Foster-Tucker, Rachel Truscon, Emileigh Johnson, Casey Juntila, Dolapo R. Raji, Lara J. Thomas, William J. Fitzsimmons, Julie Gilbert, Leigh Papalambros, Ankur Holz, Amy Callear; **University of Washington:** Zack Acker, Julia Bennett, Erica Clark, Sarah Cox, Mark Drummond, Brenna Ehmen, Collrane Frivold, Luis Gamboa, Peter Han, Alex Harteloo, Sarah Heidl, Madison Hollcroft, Kristen Huden, Melissa MacMillan, Kathryn McCaffrey, Lani Regelbrugge, Jeremy Stone, Tessa Wright.

