## Supplementary material for "Effectiveness of mRNA COVID-19 vaccines and hybrid immunity in preventing SARS-CoV-2 infection and symptomatic COVID-19 among adults in the United States": Adult_VE_Supplementary_Materials_11_1_2024.docx

#### **Supplemental Materials**

Table e4. COVID-19 vaccine effectiveness against laboratory-confirmed SARS-CoV-2 infection among adults by vaccine type, and interval since receipt of dose, November 27, 2022 – July 31, 2023

Table e5. Combined protection from vaccination and prior infection against laboratory-confirmed SARS-CoV-2 infection among adults by vaccine type and interval since receipt of dose, November 27, 2022 – July 31, 2023

### Supplemental methods:

*Time since prior infection imputation*

The date of the most recent prior SARS-CoV-2 infection was used to create a time since prior infection variable. A small percentage of participants (5.9%) were classified as having a prior infection only by a positive nucleocapsid result at enrollment and did not self-report a prior positive. For these participants time since infection was imputed.

Previous publications have shown a relationship between nucleocapsid levels and time since infection [26]. We assumed participants were at least 30 days from infection and modeled the nucleocapsid decline with repeated measures linear regression. Each model had log days since prior infection as the outcome with log nucleocapsid values and baseline nucleocapsid blood draw date as predictors. Participants who had nucleocapsid results and a self-reported prior positive date or in-study positives were included. The resulting betas from these models were then used to estimate a predicted days since infection for each participant who had a missing date of infection.

The average residual across models was 125 days, with a median of 78. This suggests the linear prediction is reasonable for creating the final time since infection variable which uses categories time in 3–6-month increments.

### Table e1. SARS-CoV-2 RT-PCR testing practices for weekly and symptomatic respiratory swabs, whole genome sequencing methods, and SARS-CoV-2 serologic testing assays by study site.

|  | CASCADIA | COVE | VIEW |
| --- | --- | --- | --- |
| Collection Media | RHINOsticTM Automated Nasal Swab (Rhinostics RH-S000001), returned in a MatrixTM 1.0 mL ScrewTop Tube (Thermo Fisher 3741) with no transport medium | Dry Swabs with incubation in DNA/RNA shield (Zymo) | Hologic Direct Load tubes |
| Collection Protocol | Self-collection and sent by courier to University of Washington Brotman Baty Institute | Self-collection and mailed to central lab | Self-collection and mailed to central lab |
| Pooling Protocol | None | 4x pool created prior to extraction. Individual samples in pools with Ct ≤ 40 are retested. | None |
| Extraction Protocol | Rehydration with TE and treatment with Proteinase K followed by heat inactivation, or extracted using Roche MagnaPure 96 Small Volume Total Nucleic Acids kit | Kingfisher MagMax VPII Nucleic Acid Isolation Kit | Hologic Integrated Panther Fusion System |
| Positive Controls | RNase P for human component in each sample, positive controls for each viral target | Internal MS2 Control | Hologic internal control |
| RT-PCR Assay | Laboratory developed multiplexed assay for Influenza A and B + SARS-CoV-2 (2 targets) + RSV A and B + RNase P | Applied Biosystems TaqMan™ SARS-CoV-2, Flu A/B, RSV RT-PCR Assay Kit | Hologic Panther platform |
| Determination of  Positive Result | >2 replicate reactions Ct <= 40 for SARS-CoV-2  2 replicate reactions < 40 for each of the other targets | Ct ≤40 | Ct ≤40 |
| Whole genome sequencing | Viral genome sequencing was attempted on all SARS-CoV-2-, influenza, RSV- positive samples with an average Ct value of ≤30 using custom molecular inversion probes from Molecular Loops [19]. Lineages meeting pre-defined quality criteria were assigned using PANGOLIN [22] | Specimens were processed for whole genomic sequencing using the ARTIC Network protocol (v5.3.2 primers) on an Illumina instrument [20, 21]. Lineages meeting pre-defined quality criteria were assigned using PANGOLIN [22] | Specimens were processed for whole genomic sequencing using the ARTIC Network protocol (v5.3.2 primers) on an Illumina instrument [20, 21]. Lineages meeting pre-defined quality criteria were assigned using PANGOLIN [22] |
| Serology sample collection | Venous blood draw or Tasso device | Venous blood draw or Tasso device | Mitra device |
| Serologic assay | MSD | MSD ^a^ | MSD ^a^ |
| Anti-nucleocapsid antibody cutoff | 5000 AU/mL | 5000 AU/mL | 5000 AU/mL |
| Sensitivity/specificity | 93.8%/100% | 93.8%/100% | 93.8%/100% |

Abbreviations: PCR = polymerase chain reaction; Ct = cycle threshold; PANGOLIN = phylogenetic assignment of named global outbreak lineages; ELISA = enzyme linked immunosorbent assay; MSD = Meso Scale Discovery.

^a^Serum collection for study participants is optional.

### Table e2. COVID-like illness (CLI) symptoms included in the weekly survey by study site.

| Covid-like illness symptoms | CASCADIA | CoVE | VIEW |
| --- | --- | --- | --- |
| Fever | X | X | X |
| Chills | X | X | X |
| Cough | X | X | X |
| Shortness of breath | X | X | X |
| Sore throat | X | X | X |
| Muscle or body aches | X | X | X |
| Headache | X | X | X |
| Change in smell or taste | X |  | X |
| Nasal congestion or runny nose | X | X | X |
| Diarrhea | X | X^a^ | X |
| Fatigue/being run-down | X | X | X |
| Persistent pain or pressure in chest | X |  |  |
| Pale, gray, or blue colored skin, lips, nail beds | X |  |  |
| Decreased appetite |  | X^a^ |  |
| Nausea/vomiting |  | X^a^ | X |

^a^Symptoms did not trigger a swab outside of the weekly cadence.

### Table e3. Characteristics of participants by study site, Sept 1, 2022 – July 31, 2023.

|  | CASCADIA  No. (Col %) | | CoVE  No. (Col %) | | VIEW  No. (Col %) | | P-value^a^ |
| --- | --- | --- | --- | --- | --- | --- | --- |
| Total (Row %) | 1,673 | 50.0% | 354 | 10.6% | 1,316 | 39.4% | 0.020 |
| Sex |  |  |  |  |  |  |  |
| Female | 1,114 | 66.6% | 228 | 64.4% | 905 | 68.8% |  |
| Male | 557 | 33.3% | 124 | 35.0% | 405 | 30.8% |  |
| Non-female or male | 0 | 0.0% | 0 | 0.0% | 5 | 0.4% |  |
| Age median (IQR) | 41.0 | 38.0 - 45.0 | 43.5 | 38.0 - 52.7 | 40.7 | 31.9 - 50.9 | <0.001 |
| Age group (years) |  |  |  |  |  |  | <0.001 |
| 18-49 | 1,673 | 100% | 244 | 68.9% | 963 | 73.2% |  |
| 50-64 | 0 | 0.0% | 78 | 22.0% | 308 | 23.4% |  |
| 65+ | 0 | 0.0% | 32 | 9.0% | 45 | 3.4% |  |
| Race/Ethnicity |  |  |  |  |  |  | <0.001­­ |
| White, non-Hispanic | 1,254 | 75.4% | 286 | 81.3% | 772 | 62.1% |  |
| Hispanic or Latino | 108 | 6.5% | 28 | 8.0% | 116 | 9.3% |  |
| Multiple races, non-Hispanic | 79 | 4.7% | 10 | 2.8% | 38 | 3.1% |  |
| Black, non-Hispanic | 38 | 2.3% | 4 | 1.1% | 275 | 22.1% |  |
| Other, non-Hispanic^b^ | 185 | 11.1% | 24 | 6.8% | 42 | 3.4% |  |
| Chronic Conditions^c^ |  |  |  |  |  |  | <0.001 |
| None | 810 | 48.4% | 155 | 43.8% | 366 | 27.8% |  |
| 1 or more | 863 | 51.6% | 199 | 56.2% | 950 | 72.2% |  |
| Individuals living in household |  |  |  |  |  |  | <0.001 |
| 1 | 22 | 1.3% | 87 | 24.6% | 166 | 12.8% |  |
| 2 | 129 | 7.7% | 47 | 13.3% | 477 | 36.7% |  |
| 3 | 448 | 26.8% | 47 | 13.3% | 260 | 20.0% |  |
| ≥4 | 1,073 | 64.2% | 173 | 48.9% | 396 | 30.5% |  |
| Weekly swab adherence (%), median (IQR) | 89.6 % | 81.3 – 95.5 | 90.9 % | 79.3 – 97.8 | 75.9 % | 66.7 - 85.2 | <0.001 |
| Prior infection^d^ | 639 | 38.2% | 179 | 19.8% | 644 | 48.9% | <0.001 |
| SARS-CoV-2 infection during the study | 383 | 22.9% | 70 | 19.8% | 274 | 20.8% | 0.251 |
| Symptomatic COVID-19^e^ | 315 | 82.2% | 48 | 68.6% | 120 | 43.8% | <0.001 |

Abbreviations**:** IQR = interquartile range.

^a^Fisher’s exact tests, Kruskal-Wallis rank sum test, and Pearson’s Chi-squared tests were used to calculate p-values.

^b^The ‘Other, non-Hispanic’ category includes participations who identified as American Indian non-Hispanic, Alaska Native non-Hispanic, Asian non-Hispanic, Black and African American non-Hispanic, and Native Hawaiian/Pacific Islander non-Hispanic.

^c^Chronic conditions for CASCADIA and CoVE included: asthma, heart disease, sleep apnea, down syndrome, diabetes, cancer, autoimmune disease, liver disease, kidney disease, hematological disease, neurologic or neuromuscular disease, stroke, deep vein thrombosis or pulmonary embolism, anxiety, depression, immunosuppression, hypertension and thyroid disease. For VIEW: asthma, chronic pulmonary disease, obesity, heart disease, diabetes, liver disease, kidney disease, cancer, arthritis, hematological disease, neurologic or neuromuscular disease, stroke, deep vein thrombosis or pulmonary embolism, anxiety, depression, immunosuppression, hypertension and thyroid disease.

^d^Prior infection was defined as laboratory-confirmation of infection by RT-PCR from a study-collected specimen, positive anti-N SARS-CoV-2 antibody, or self-report of infection prior to enrollment or September 1, 2022.

^e^Symptomatic COVID-19 was defined as those with a positive RT-PCR test and at least two COVID-like illness symptoms reported within seven days of the specimen collection date.

##### Table e4. COVID-19 vaccine effectiveness against laboratory-confirmed SARS-CoV-2 infection among adults by vaccine type, and interval since receipt of dose, November 27, 2022 – July 31, 2023

| Interval since receipt of dose | Contributing participants^a^ | Median observation day from vaccination^b^ (IQR) | SARS-CoV-2 infections | Crude incidence rate of SARS-CoV-2 infections per 1000 PD | Unadjusted VE (95% CI) | Adjusted VE against infection (95% CI) |
| --- | --- | --- | --- | --- | --- | --- |
| Unvaccinated or monovalent vaccine ≥180 days ago | 1644 | 479 (335, 586) | 311 | 1.72 (1.54, 1.93) | REF | REF |
| Monovalent vaccine, <180 days ago | 376 | 117 (85, 150) | 55 | 1.59 (1.20, 2.07) | 38.5 (14.8, 55.6) | 24.2 (-6.5, 46.9) |
| Bivalent vaccine, 7-60 days ago^c^ | 708 | 40 (27, 51) | 24 | 1.03 (0.66, 1.53) | 58.6 (35.9, 73.3) | 46.2 (17.5, 66.3) |
| Bivalent vaccine, 7-179 days ago | 1426 | 112 (75, 147) | 189 | 1.34 (1.15, 1.54) | 43.8 (30.5, 54.6) | 26.9 (9.6, 40.6) |
| Bivalent vaccine ≥180 days ago | 1396 | 236 (207, 266) | 119 | 0.96 (0.80, 1.15) | 23.6 (2.8, 39.9) | 8.5 (-19.0, 31.8) |
| Bivalent vaccine, overall | 1720 | 171 (108, 231) | 308 | 1.16 (1.04, 1.30) | 36.9 (24.8, 47.0) | 19.9 (3.2, 33.2) |

Abbreviations**:** IQR = interquartile range; PD = person-days; Ref = referent group; VE = vaccine effectiveness.

^a^Contributing participants in vaccination categories do not equal the number of participants in the study because participants could contribute to more than one vaccination category since vaccination status is time-varying.

^b^Adjusted estimates control for coefficient estimates of age, sex, race/ethnicity, underlying health conditions, prior infection status, geographic site, household size, and 7-day average of COVID-19 cases per 100 000 by site.

##### Table e5. Combined protection from vaccination and prior infection against laboratory-confirmed SARS-CoV-2 infection among adults by vaccine type and interval since receipt of dose, November 27, 2022 – July 31, 2023

| Vaccine type, interval since receipt of vaccine, and prior infection status | Contributing participants^a^ | Median observation day from vaccination^b^ (IQR) | SARS-CoV-2 infections | Crude incidence rate of SARS-CoV-2 infections per 1000 PD | Unadjusted VE (95% CI) | Adjusted VE against infection^c^ (95% CI) | Symptomatic SARS-CoV-2 infections^d^ | Crude incidence rate of symptomatic COVID-19 Per 1000 PD | Unadjusted VE against symptomatic COVID-19  (95% CI) | Adjusted VE against symptomatic COVID-19  (95% CI) |
| --- | --- | --- | --- | --- | --- | --- | --- | --- | --- | --- |
| Naive^c^ & unvaccinated or monovalent vaccine ≥180 days ago | 804 | 474 (297, 582) | 172 | 2.01 (1.72, 2.33) | REF | REF | 94 | 1.10 (0.89, 1.34) | REF | REF |
| Prior infection^c^ & monovalent vaccine ≥180 days ago | 738 | 484 (369, 590) | 119 | 1.42 (1.17, 1.70) | 31.1 (12.7, 45.7) | 28.6 (8.6, 43.7) | 40 | 0.48 (0.34, 0.65) | 57.7 (37.9, 71.1) | 56.4 (39.3, 71.4) |
| Prior infection^c^ & bivalent vaccine ≥180 days ago | 610 | 232.5 (205, 264) | 37 | 0.71 (0.50, 0.97) | 53.6 (32.1, 68.4) | 40.9 (14.7, 61.3) | 21 | 0.40 (0.25, 0.61) | 55.8 (25.2, 73.8) | 56.7 (30.5, 77.0) |
| Prior infection^c^ & monovalent vaccine <180 days ago | 142 | 117 (83, 149) | 13 | 0.92 (0.49, 1.57) | 68.5 (43.1, 82.6) | 60.0 (25.9, 82.1) | 6 | 0.42 (0.16, 0.92) | 72.0 (34.5, 88.1) | 80.6 (52.4, 93.7) |
| Prior infection^c^ & bivalent vaccine <180 days ago | 606 | 110 (72, 146) | 41 | 0.62 (0.44, 0.84) | 77.4 (67.7, 84.2) | 65.5 (49.5, 77.0) | 21 | 0.32 (0.20, 0.48) | 77.3 (62.6, 86.2) | 73.0 (62.2, 87.1) |

Abbreviations**:** IQR = interquartile range; PD = person-days; Ref = referent group; VE = vaccine effectiveness.

^a^Contributing participants in vaccination categories do not equal the number of participants in the study because participants could contribute to more than one vaccination category since vaccination status is time-varying.

^b^Adjusted estimates control for coefficient estimates of age, sex, race/ethnicity, underlying health conditions, prior infection status, geographic site, household size, and 7-day average of COVID-19 cases per 100 000 by site.

^c^Naive is defined as no evidence of prior infection before September 1, 2022. Prior infection was defined as laboratory-confirmation of infection by RT-PCR from a study-collected specimen, positive anti-N SARS-CoV-2 antibody, or self-report of infection prior to enrollment or September 1, 2022 (whichever occurred later).

### Figure e1. Whole genome sequencing results for a subset of laboratory-confirmed SARS-CoV-2 respiratory specimens, September 1, 2022 – July 31, 2023.

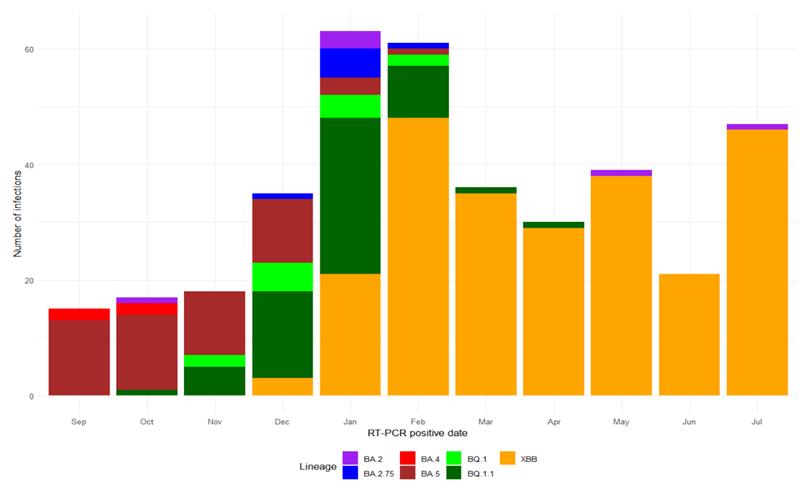
